# Effect of real-time feedback-based core stabilization training using a sling on balance and gait in patients with stroke: Randomized Controlled Trial

**DOI:** 10.1101/2023.11.17.23298709

**Authors:** Ja-young Yoo, Jungae An, Byounghee Lee

**Affiliations:** Graduate School of Physical Therapy, Sahmyook University, Seoul 01795, Korea; Department of Physical Therapy, Sahmyook University, Seoul 01795, Korea

**Keywords:** stroke, real-time feedback, core, balance, gait

## Abstract

**Background:** Balance impairments commonly occur in patients after stroke. Research is warranted to improve the efficiency of rehabilitation by combining core stabilization training, such as trunk exercises, and real-time feedback. This study aimed to evaluate the effect of real-time feedback-based core stabilization training (RFCST) using a sling on the dynamic balance and gait of patients with stroke.

**Methods:** Thirty-eight patients with stroke were randomly assigned to either RFCST using a sling group (*n=* 19) or a control group (*n=* 19). Each group was trained for 30 min daily, 3 times a week for 4 weeks. The Trunk Impairment Scale (TIS), Functional Reach Test (FRT), Postural Assessment Scale for Stroke (PASS), Timed Up and Go (TUG) test, and gait parameters were assessed using the GAITRite system before and after the intervention.

**Results:** The results showed a significant interaction between Group*Time effect F(1, 36)= 36.068, *p*<0.001, η²_p_= 0.5 in TIS; F(1, 36)= 63.890, *p*<0.001, η²_p_= 0.640 in FRT; F(1, 36)= 89.283, *p*<0.001, η²_p_= 0.713 in PASS, F(1, 36)= 150.893, *p*<0.001, η²_p_= 0.807 in TUG; F(1, 36)= 27.275, *p*<0.001, η²_p_= 0.431 in gait velocity; F(1, 36)= 54.401, *p*<0.001, η²_p_= 0.447 in cadence; F(1, 36)= 5.601, *p*=0.023, η²_p_= 0.135 in step length; F(1, 36)= 22.559, *p*<0.001, η²_p_ = 0.385 in stride length; F(1, 36)= 15.516, *p*<0.001, η²_p_ = 0.301 in swing phase rate; and F(1, 36)= 28.451, *p*<0.001, η²_p_ = 0.441 in stance phase rate.

**Conclusion:** Based on these results, it can be concluded that RFCST using a sling can improve dynamic balance and gait parameters in patients with stroke.

## 1. Introduction

A stroke occurs when a blood vessel in the brain becomes blocked or bursts, leading to acute neurological deficits caused by damage to the central nervous system (1). Stroke survivors may experience muscle weakness, abnormal muscle tone and movement patterns, asymmetric body balance, and difficulty in walking and balance. Moreover, there may be deficits in the ability to transfer weight, resulting in disability, extremity dysfunction, and difficulty in attaining dynamic postures such as walking and exercising (2, 3).

In patients with stroke, trunk dysfunction includes decreased sitting balance, decreased coordination, decreased trunk and lower extremity muscle strength, and lack of trunk position detection ability (4). Rehabilitation programs, including trunk stability training, have been reported to improve static and dynamic trunk balance (5). Proactive postural control of the trunk occurs before limb movement in humans (6), and impairment of trunk control leads to reduced trunk movement, which is an appropriate response to reduced pelvic movement and loss of balance(7). A study on the relationship between trunk and limb muscle dysfunction reported that distance, speed, and weight-bearing ability of the lower extremities improved by improving the trunk control ability (8). Comparison of the gaits of hemiplegic patients due to stroke and normal adults revealed that the symmetry of trunk movement was reduced, which in turn correlated with gait speed (9). Therefore, effective trunk rehabilitation interventions to improve balance and walking abilities in patients with stroke are receiving increasing attention (10).

The sling exercise uses a rope suspended from the ceiling to reduce the body load for the patient’s active exercise. The hanging rope has an unstable support surface; therefore, it has the advantage of simultaneously contracting the agonist, antagonist, and synergistic muscles through stimulation of the neuromuscular system (11, 12). Core muscles generate and control all the forces and movements of the human body. If these muscles are stretched and strengthened in patients with stroke, the mobility and stability of posture would increase to control and balance the body (12). Core stabilization exercises promote an integrated system from the toes to the torso by continuous segmental movements that promote the ability of the body’s load to translocate across the lumbar and sacral vertebrae (13). Of the various types of core stabilization exercises, exercises performed on an unstable surface increase neural action on muscles and identification of motor units. Moreover, they also result in increased activity of synergistic action of muscles, which in turn increases the stability around the compound joints and muscle strength because more muscles can be mobilized (12). Therefore, to improve the trunk control ability of patients with stroke, exercise programs that strengthen the core muscles, which are the central muscles of the body, are necessary (14). Balance training using visual and auditory feedback has been studied as a method to improve the balance control ability of patients with hemiplegia due to stroke (15–17). Real-time visual feedback provides real-time visual information during movement so that patients can identify the position and location of their center of gravity during postural changes. Moreover, it enables patients to acquire their posture information, which can be used for the maintenance of control and posture (18, 19). This leads to high levels of motivation, increased compliance, and low training consumption, which has a positive impact on rehabilitation training outcomes (20, 21). However, despite these effects, studies combining real-time feedback with core stabilization exercises using a sling are scarce. Therefore, this study aimed to investigate the effects of core stabilization training combined with real-time feedback on dynamic balance ability and gait in patients with stroke.

## 2. Materials and Methods

### 2.1. Study participants

This study included 38 adult patients who had suffered a stroke and were admitted to a rehabilitation hospital. The sample size of this study was calculated using G*Power Version 3.1.9.7 (Franz Faul, University Kiel, Germany, 2020). Based on the results of the independent sample t-test, which served as the primary analytical method for evaluating the program’s effect, a sample size of 34 was deemed necessary to maintain an effect size of 0.5, a significance level (α) of 0.05, and a power of 0.8 in a two-tailed test. Therefore, we recruited a total of 40 patients with stroke for this study, taking into consideration the possible dropouts during the study duration. The inclusion criteria were: hemiplegic patients aged 40-75 years who had been diagnosed with stroke at least six months, those who scored more than 24 points on the Mini-Mental State Examination-K, and walked more than 10 m using a mono-cane. The exclusion criteria were: patients with the other neurologic or orthopedic disorder; those with visual problems and hemianopia, and those who could not evaluate because they could not understand verbal cues such as cognitive impairment. All participants were explained the purpose of the study and the necessary aspects, and the study was conducted after obtaining a written informed consent from participants or their caregivers. The recruitment period for this study was from July 1, 2021 to August 29, 2021. This study was approved by the SAHMYOOK UNIVERSITY Institutional Review Board (SYUIRB2-1040781-A-N-012021059HR) and the Clinical Research Information Service (https://cris.nih.go.kr/cris/index/index.do, Registration Number: KCT0006552). The rights of participants were protected according to the ethical principles of the Declaration of Helsinki.

**Figure 1.**
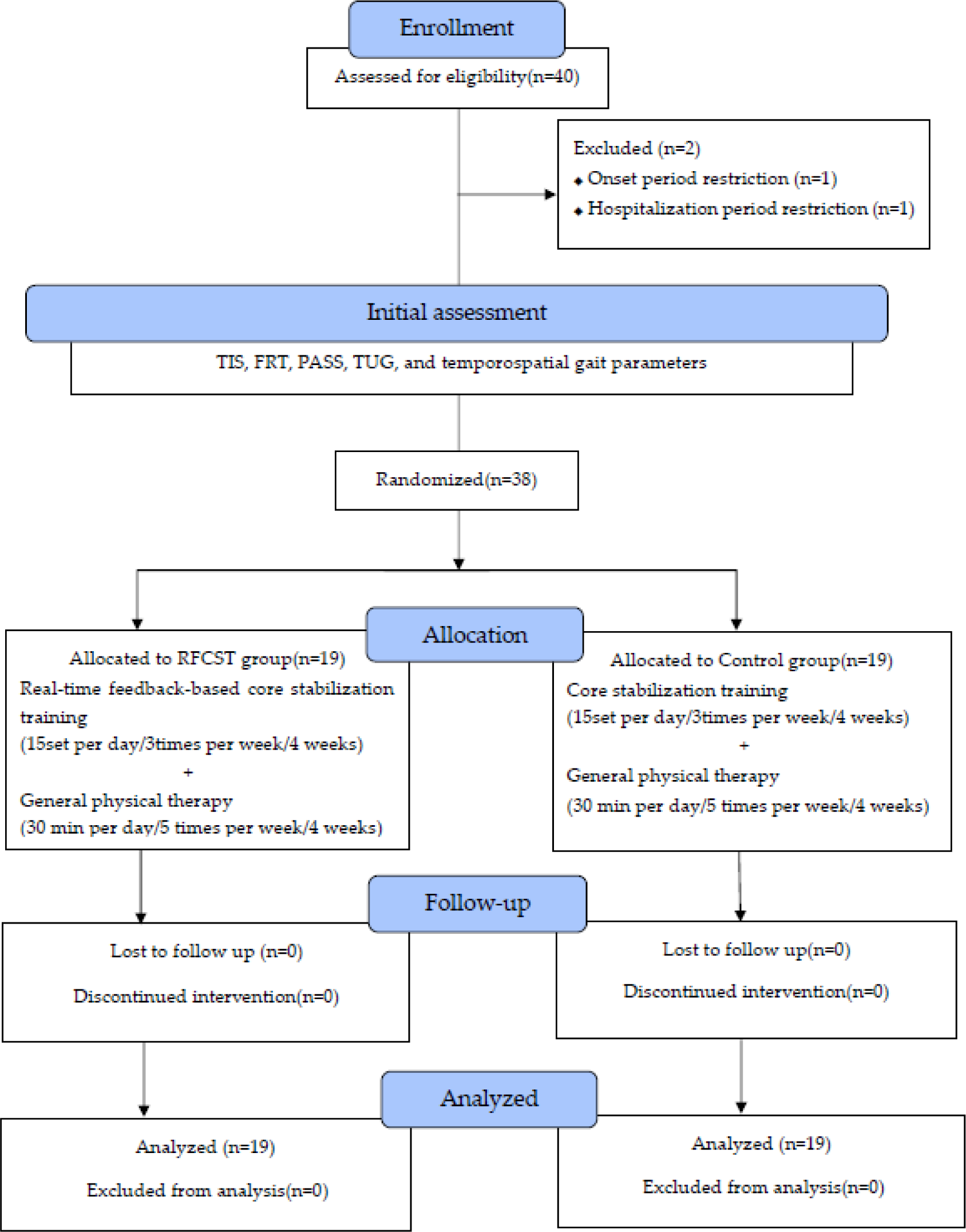
Flow chart demonstrating the patient flow and study procedure

### 2.2. Experimental Procedures

This study was conducted using a pretest-posttest control group design. Two patients who did not meet the study selection criteria were excluded. One patient was eliminated due to onset period restriction and the other due to hospitalization period restriction. To minimize selection bias, Research Randomizer (http://www.randomizer.org, accessed on August 1, 2021) was used. A research nurse who was not involved in this study used a computerized random generator to assign participants randomly. Following the initial assessment, 38 patients who agreed to participate and met the eligibility criteria were divided into either the experimental an experimental group (n = 19) that underwent real-time feedback-based core stability training (RFCST) or a control group (n = 19) that underwent core stability training. The allocation was done by allowing each participant to select one of two concealed envelopes containing the group assignments.

The real-time feedback-based training group underwent real-time feedback-based sling core stabilization training, whereas the control group underwent core stabilization training using a sling without feedback. The training was applied three times a week for 30 min each, and after 4 weeks, dynamic balance and walking ability were evaluated using the same measurement tool. Both groups received general exercise therapy, including the central nervous system approach for 30 min, 5 times a week.

#### 2.2.1. Real-time feedback

The stroke patient was allowed to adjust the height randomly using an elastic cord while supporting the pelvis on a wide sling in the supine position. The patients were instructed to raise their pelvis to a point marked by the physiotherapist on the wall using a laser point. Real-time feedback was provided verbally and visually by the patient. This helped the patients maintain the posture of lifting the pelvis to the point marked on the wall (22).

##### Core stabilization training with a sling

Core stabilization training was performed using a sling exercise program for 30 min, 3 times a week for 4 weeks. The training was performed using a sling device while lying on a height-adjustable Bobath table, and the level of difficulty was adjusted by parking. In the first and second weeks of training, three sets of five repetitions were provided, with 60 s of rest between the sets. In the third week of training, the red elastic cord was increased, and three sets of five repetitions with 60 seconds of rest between sets were provided. In the fourth week of training, the black elastic cord was lowered to provide three sets of five repetitions, with a rest period of 60 s between the sets (23) (Table 1).

**Table 1.** Core stabilization training using a sling.

| Week | Core stabilization training | Times |
| --- | --- | --- |
| 1<sup>st</sup> and 2<sup>nd</sup> week | 1. In a supine position, support the pelvis with a wide sling, the healthy leg with the other narrow sling, and hold the knee at 90° at ankle height for 10 seconds.<br>2. After changing the wide sling to a led elastic cord to support the pelvis, the height is adjusted randomly and maintained as high as possible. | 5 times,<br>3 sets |
| 3<sup>rd</sup> week | 1. Adjust the posture with the trunk, lower limbs, and arms on the Bobath table, support the pelvis with a wide sling and the affected leg with a red (strong) elastic cord on a narrow sling, and press it down for 10 seconds.<br>2. With the affected leg supported by an unstable, lift the unaffected leg in the air and hold it for 10 seconds. | 5 times,<br>3 sets |
| 4<sup>th</sup> week | 1. Adjust the posture with the trunk and lower limbs while placing the arms on the abdomen without placing them on the Bobath table. After supporting the pelvis with a wide sling, support the affected leg with a black (weak) elastic cord on a narrow sling and press it down. This position is to be held for 10 seconds.<br>2. Support the affected leg as unstable. Hold the unaffected leg in the air for 10 seconds. | 5 times,<br>3 sets |
| Total training time was 30 min, with a rest time of 60 s between the sets. |  |  |

##### General physical therapy

General exercise therapy is a one-on-one treatment provided by a physical therapist. Neurologic facilitation approaches such as Bobath’s neurodevelopmental therapy and proprioceptive neuromuscular facilitation (PNF) were performed once a day for 30 min, five times for four weeks.

### 2.3. Outcome Measurements

#### 2.3.1. Primary Outcomes: Dynamic balance

The trunk impairment scale (TIS), functional reach test (FRT), and postural assessment scale for stroke (PASS) were used to assess dynamic balance. All the evaluations were conducted by a physical therapist who had more than 10 years of clinical experience and did not treat the participants. Measurements were repeated three times, two days before and two days after the training by the same examiner to minimize measurement errors in an independent laboratory. The average values were used.

In TIS, the patient was instructed not to lean on the edge of the mat or treatment table or support it with his or her hands. Further, the patients were instructed to lie down on the floor with thighs being in complete contact with the mat or table and feet being as wide apart as the hips. The test commenced with knees bent at 90 degrees and arms resting on the legs. The TIS comprises three items and ranges from a minimum score of 0 to a maximum of 23 points, it consists of static sitting posture balance ability (7 points) and evaluates whether an individual can maintain balance using the non-affected lower extremity crossed over the paralyzed lower extremity in a sitting position, dynamic sitting balance ability evaluates the movement of separating the upper and lower parts of the trunk through lateral flexion in a sitting position (14 points), coordination ability (6 points), which evaluates the horizontal rotational movement of the shoulder and pelvic girdle in a sitting position, with higher scores indicating better trunk control ability (24). TIS was found to have a high intraclass correlation coefficient (ICC=0.96) for test-retest reliability (25). To measure static and dynamic sitting balance, reliability, efficacy, and internal consistency (Cronbach’s α=0.89) were reported for patients with stroke (26).

The functional reach test does not require any equipment and can be easily employed clinically (27). It refers to the maximum distance that can be reached forward beyond the length of the arm by extending the arm while maintaining it without moving the foot in a standing position. This balance test has been widely used to predict falls due to its high reliability and validity (28). It has been reported the arm’s reach increases with an increase in the size of the trunk and arm movements, and that the trunk plays a crucial role in overall movement (29).

The PASS was used to evaluate postural control performance in patients with stroke, it was developed by modifying and supplementing the Fugl-Meyer Assessment – Balance (FM-B)(30). Moreover, it is a useful clinical tool for diagnosing the condition of patients with stroke because it can be evaluated easily and within a short duration of 1-10 min (31). PASS comprises three basic postures: lying, sitting, and standing. It consists of 12 items, including five posture maintenance items and seven posture change items. Dynamic balance ability was evaluated as good. PASS has reportedly shown high reliability and validity in patients with stroke (ICC = 0.97; inter-rater reliability, r = 0.98) (32).

#### 2.3.2. Secondary Outcomes: Gait ability

Spatiotemporal variables were evaluated using a timed up-and-go (TUG) test and GaitRite to examine walking ability. TUG test assesses functional mobility and dynamic balance. It was used to predict the risk of falls by evaluating the balance ability and functional movement of an older individual. This test has recently been used not only for older individuals at high risk of falls, but also for patients with stroke, Parkinson’s disease, and arthritis. It can act as a factor in determining the patient’s walking speed, falls, and community walking ability (33). The time taken to return to the chair by going back and forth 3 m after standing up at the command “start” while sitting in a chair with armrests was measured. The patients could use the shoes and aid that they usually wore during the measurement but without the help of the therapist. In this method, the intra-rater reliability was r=0.99, and the inter-rater reliability was r=0.98, which was high (34). It is highly valid for evaluating balance, walking speed, and functional movements (35).

The gait ability test measures temporal and spatial gait ability using a gait analyzer (GaitRite, CIR System Inc., USA, 2008) to collect data for quantitative gait analysis of the patient’s gait type. The gait analyzer (GaitRite) is an electronic gait board measuring 5 m in length, 0.6 cm in height, and 61 cm in width. It consists of 16,128 sensors with a diameter of 1 cm that are vertically arranged at intervals of 1.27 cm along the walking board to collect information on temporal and spatial variables. Information on the collected temporal and spatial variables was processed using GaitRite GOLD, Version 3.2b (CIR System Inc., USA, 2007) software. The experimental method involved the participant standing in front of a walking board, and when the examiner sent a verbal signal, they walked at the most comfortable speed to move out of the gait board. Gait speed, cadence, step length, stride length, and spatial gait characteristics of single limb support percentage were recorded through informational analysis. The participants were given a 10-minute break between measurements to minimize fatigue. The reliability of this test was r=0.90, and the correlation coefficient within all gait measures of comfortable gait speed was > 0.96 (36).

### 2.4 Statistical analysis

All tasks and statistical analyses were performed using SPSS ver. 22.0(IBM, Chicago, IL, USA). Data were presented as mean and standard deviation. All participants were subjected to Kolmogorov-Smirnov normality verification, and as a result, they were normally distributed. Descriptive statistics were used to analyze the general characteristics of the participants. Moreover, homogeneity tests were performed before the experiment by the various variables of the two groups. To determine the differences between groups, the experimental results were analyzed using an independent sample t-test. A paired-sample t-test was used to compare before and after training data within the group. The interaction effect between groups over time was analyzed using a two-way repeated measures analysis of variance (ANOVA). The level of significance was set at 0.05.

## 3. Results

### 3.1. General and Clinical Characteristics of the Study Participants

The general and clinical characteristics along with the results of homogeneity tests of the participants are presented in Table 2.

**Table 2.**
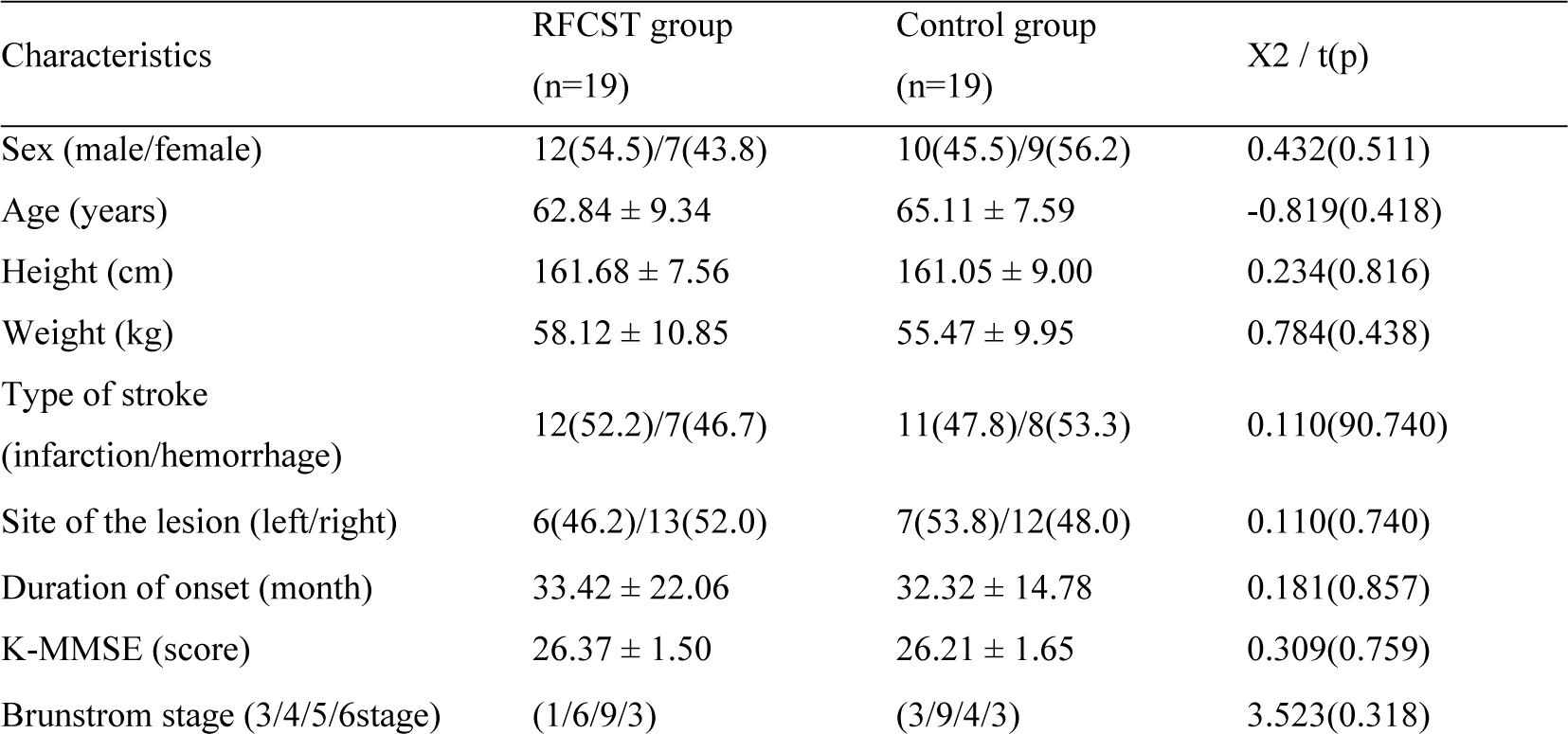
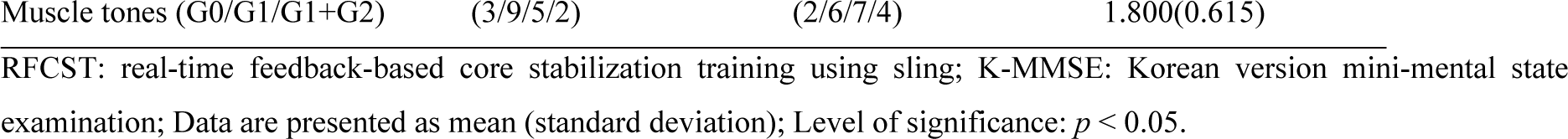
General and clinical characteristics of the study participants *(N*=38)

### 3.2. Changes in dynamic balance

The group that underwent real-time feedback-based core stabilization training using a sling demonstrated a significant improvement in TIS, FRT, and PASS compared to the control group (*p* < 0.05). The results revealed a significant interaction between Group × Time, with F-values of F(1, 36) = 36.068, *p* < 0.001, η²_p_ = 0.5 for TIS; F(1, 36) = 63.890, *p* < 0.001, η²_p_ = 0.640 for FRT; and F(1, 36) = 89.283, *p* < 0.001, η²_p_ = 0.713 for PASS (as shown in Table 3). When examining the group effect, only PASS showed a significant difference (p<0.05).

**Table 3.**
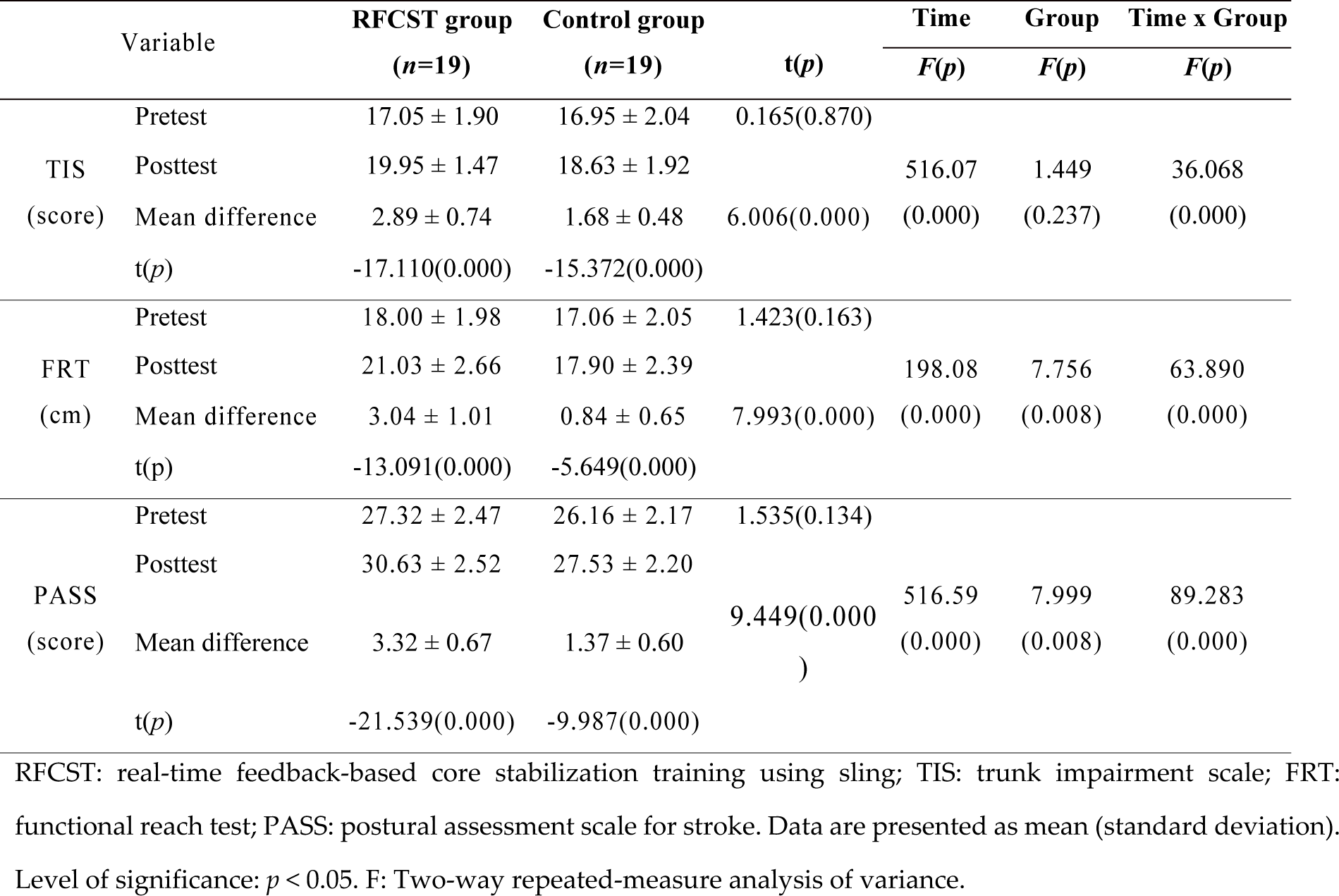
Changes in dynamic balancing capacity according to the experimental methods (*N*=38)

| Variable |  | RFCST group<br>(n=19) | Control group<br>(n=19) | t(p) | Time<br>F(p) | Group<br>F(p) | Time x Group<br>F(p) |
| --- | --- | --- | --- | --- | --- | --- | --- |
| TIS<br>(score) | Pretest | 17.05 ± 1.90 | 16.95 ± 2.04 | 0.165(0.870) | | | |
| | Posttest | 19.95 ± 1.47 | 18.63 ± 1.92 | | 516.07 | 1.449 | 36.068 |
| | Mean difference | 2.89 ± 0.74 | 1.68 ± 0.48 | 6.006(0.000) | (0.000) | (0.237) | (0.000) |
|  | t(p) | -17.110(0.000) | -15.372(0.000) |  |  |  |  |
| FRT<br>(cm) | Pretest | 18.00 ± 1.98 | 17.06 ± 2.05 | 1.423(0.163) | | | |
| | Posttest | 21.03 ± 2.66 | 17.90 ± 2.39 | | 198.08 | 7.756 | 63.890 |
| | Mean difference | 3.04 ± 1.01 | 0.84 ± 0.65 | 7.993(0.000) | (0.000) | (0.008) | (0.000) |
|  | t(p) | -13.091(0.000) | -5.649(0.000) |  |  |  |  |
| PASS<br>(score) | Pretest | 27.32 ± 2.47 | 26.16 ± 2.17 | 1.535(0.134) | | | |
| | Posttest | 30.63 ± 2.52 | 27.53 ± 2.20 | | 516.59 | 7.999 | 89.283 |
| | Mean difference | 3.32 ± 0.67 | 1.37 ± 0.60 | 9.449(0.000) | (0.000) | (0.008) | (0.000) |
|  | t(p) | -21.539(0.000) | -9.987(0.000) |  |  |  |  |

RFCST: real-time feedback-based core stabilization training using sling; TIS: trunk impairment scale; FRT: functional reach test; PASS: postural assessment scale for stroke. Data are presented as mean (standard deviation). Level of significance: $p < 0.05$ . F: Two-way repeated-measure analysis of variance.

### 3.3. Changes in temporal gait parameters

The temporal gait parameters showed a significant interaction between Group × Time effect, with F values of F(1, 36)= 150.893, *p*<0.001, η²_p_= 0.807 in TUG; F(1, 36)= 27.275, *p*<0.001, η²_p_= 0.431 in gait velocity; and F(1, 36)= 54.401, *p*<0.001, η²_p_= 0.447 in cadence (Table 4). However, no significant group effects were observed.

**Table 4.** Changes in temporal walking capacity according to the experimental methods (*N*=38)

| Variable | | RFCST group<br>($n=19$) | Control group<br>($n=19$) | $t(p)$ | Time<br>$F(p)$ | Group<br>$F(p)$ | Time x Group<br>$F(p)$ |
| --- | --- | --- | --- | --- | --- | --- | --- |
| TUG(sec) | Pretest | 22.70 ± 7.06<sup>a</sup> | 23.76 ± 9.01 | -0.401(0.691) | | | |
| | Posttest | 20.01 ± 7.02 | 23.17 ± 8.86 | | 365.52 | 0.652 | 150.90 |
| | Mean difference | -2.70 ± 0.63 | -0.59 ± 0.40 | 12.28(0.000) | (0.000) | (0.425) | (0.000) |
| | $t(p)$ | 18.633(0.000) | 6.347(0.000) | | | | |
| Gait velocity<br>(cm/sec) | Pretest | 37.05 ± 22.80 | 31.99 ± 16.57 | 0.783(0.439) | | | |
| | Posttest | 42.62 ± 23.33 | 33.35 ± 16.77 | | 74.103 | 1.209 | 27.275 |
| | Mean difference | 5.57 ± 2.36 | 1.36 ± 2.60 | 5.223(0.000) | (0.000) | (0.279) | (0.000) |
| | $t(p)$ | -10.276(0.000) | -2.289(0.034) | | | | |
| Cadence<br>(steps/min) | Pretest | 60.76 ± 22.47 | 64.89 ± 27.64 | -0.505(0.617) | | | |
| | Posttest | 64.68 ± 21.96 | 65.93 ± 27.27 | | 50.391 | 0.090 | 29.041 |
| | Mean difference | 3.92 ± 1.68 | 1.04 ± 1.51 | 5.565(0.000) | (0.000) | (0.765) | (0.000) |
| | $t(p)$ | -10.187(0.000) | -3.015(0.007) | | | | |

RFCST: real-time feedback-based core stabilization training using sling; TUG: timed up and go test. Data are presented as mean (standard deviation). Level of significance: $p < 0.05$ . F: Two-way repeated-measure analysis of variance.

### 3.4. Changes in spatial gait parameters

The result of spatial gait parameters showed a significant interaction between Group × Time effect with F values of F(1, 36)= 5.601, *p*=0.023, η²_p_= 0.135 in step length of affected side; F(1, 36)= 22.559, *p*<0.001, η²_p_ = 0.385 in stride length of affected side; F(1, 36)= 15.516, *p*<0.001, η²_p_ = 0.301 in swing phase rate; F(1, 36)= 28.451, *p*<0.001, η²_p_ = 0.441 in stance phase rate; F(1, 36)= 34.221, *p*<0.001, η²_p_= 0.487 in single support rate; and F(1, 36)= 22.118, *p*<0.001, η²_p_= 0.381 in single support rate. The RFCST group demonstrated a significant difference in terms of all spatial gait parameters in comparison to the control group (p < 0.05).

**Table 5.** Change in spatial gait parameters according to the experimental method (*N*=38)

| Variable |  | RFCST group<br>(n=19) | Control group<br>(n=19) | t(p) | Time<br>F(p) | Group<br>F(p) | Time x Group<br>F(p) |
| --- | --- | --- | --- | --- | --- | --- | --- |
| Affected side<br>step length(cm) | Pretest | 30.52± 11.98 | 30.95 ± 6.51 | -0.138(0.892) | | | |
| | Posttest | 33.66 ± 13.81 | 32.14 ± 6.81 | | 21.785 | 0.036 | 5.601 |
| | Mean difference | 3.14 ± 2.82 | 1.18 ± 2.36 | 2.319(0.026) | (0.000) | (0.851) | (0.023) |
|  | t(p) | -4.860(0.000) | -2.189(0.042) |  |  |  |  |
| Affected side<br>stride length<br>(cm) | Pretest | 64.87 ± 27.74 | 58.06 ± 16.28 | 0.923(0.362) | | | |
| | Posttest | 69.09 ± 28.71 | 59.00 ± 16.18 | | 55.733 | 1.284 | 22.559 |
| | Mean difference | 4.23 ± 2.45 | 0.94 ± 1.76 | 4.750(0.000) | (0.000) | (0.265) | (0.000) |
|  | t(p) | -7.512(0.000) | -2.333(0.031) |  |  |  |  |
| Affected side<br>stance rate<br>(%) | Pretest | 71.50 ± 7.57 | 71.90 ± 10.41 | -0.135(0.893) | | | |
| | Posttest | 67.94 ± 7.32 | 70.99 ± 10.41 | | 80.924 | 0.348 | 28.451 |
| | Mean difference | -3.56 ± 1.91 | -0.91 ± 1.03 | -5.334(0.000) | (0.000) | (0.559) | (0.000) |
|  | t(p) | 8.147(0.000) | 3.847(0.001) |  |  |  |  |
| Affected side<br>single support<br>rate(%) | Pretest | 23.71 ± 9.59 | 22.51 ± 9.40 | 0.391(0.698) | | | |
| | Posttest | 26.33 ± 9.63 | 23.15 ± 9.45 | | 93.048 | 0.507 | 34.221 |
| | Mean difference | 2.62 ± 0.94 | 0.64 ± 1.13 | 5.850(0.000) | (0.000) | (0.481) | (0.000) |
|  | t(p) | -12.121(0.000) | -2.468(0.024) |  |  |  |  |
| Double support<br>rate(%) | Pretest | 47.36±15.93 | 49.23±16.67 | -0.354(0.725) | | | |
| | Posttest | 44.09±16.43 | 48.27±17.25 | | 52.803 | 0.344 | 22.118 |
| | Mean difference | -3.27±1.43 | -0.96±1.57 | -4.740(0.000) | (0.000) | (0.561) | (0.000) |
|  | t(p) | 9.975(0.000) | 2.680(0.015) |  |  |  |  |
| Affected side<br>swing rate(%) | Pretest | 28.51±7.57 | 28.10±10.42 | 0.137(0.892) | | | |
| | Posttest | 32.52±6.70 | 29.11±10.70 | | 40.423 | 0.448 | 15.516 |
| | Mean difference | 4.01±2.75 | 1.01±1.96 | 3.873(0.000) | (0.000) | (0.508) | (0.000) |
|  | t(p) | -6.352(0.000) | -2.252(0.037) |  |  |  |  |

RFCST: real-time feedback-based core stabilization training using a sling. Data are presented as mean (standard deviation). Level of significance: $p < 0.05$ . F: Two-way repeated-measure analysis of variance.

## 4. Discussion

This study aimed to explore the effects of a four-week intervention involving real-time feedback-based sling core stabilization training on dynamic balance and walking ability of 38 patients with stroke.

### 4.1. The effects of dynamic balance

The sense of trunk position is important as it provides information on trunk alignment related to gravity (37). Trunk control and dynamic balance ability are important for independent living and performing activities of daily living (ADL), such as combing, dressing, or going to the bathroom (38). Balance in a sitting position after a stroke is the most important factor in the rehabilitation effect, such as maintaining the center of the trunk by adjusting the lower trunk, pelvis, and lower extremities. This is the first goal to be achieved when treatment is being administered for neurological recovery. The second goal is to minimize activity limitations and improve walking ability to improve the quality of life (39). In this study, changes in dynamic balance before and after training were investigated using TIS, FRT, and PASS. In the TIS, real-time feedback and core stabilization training using a sling were performed for 30 min/day, three times/week for four weeks. A significant increase of 1.68 points from 16.95 to 18.63 was observed, and there was a significant difference between the groups (*p*<0.05). In a study of 32 patients with stroke, the experimental group that applied core stabilization training for 1 h, five times a week for four weeks, reported a significant increase in TIS by 2.94 points compared to the control group (*p*<0.01), which provides evidence that interventions targeting trunk muscle activation based on core stability theory have positive effects on patients with stroke (40). In this study, the RFCST group showed a significant increase of 3.03 cm from 18.00 cm before the experiment to 21.03 cm after the experiment in the FRT. In the control group, it increased significantly by 0.84 cm from 17.06 cm to 17.90 cm, and the difference between the two groups was statistically significant (*p*<0.05). In addition, in the PASS test, the RFCST group showed a significant increase of 3.31 points from 27.32 points before the experiment to 30.63 points after the experiment (*p*<0.05). A previous study wherein additional core stabilization training was performed in 1h sessions over five weeks for 110 patients with stroke in the experimental group and 110 individuals in the control group reported improvement in scores of PASS, TIS, Berg Balance Scale(BBS), and Barthel Index(BI) in the experimental group (41). These results were in concordance with our findings. In a study of 13 patients with stroke, biofeedback treatment was performed using a wearable device that affected motor learning and patient participation and reported an increase in postural maintenance activities required for dynamic balance and walking (42). In addition, as a result of applying balance training using real-time feedback to 15 patients with stroke for 20 min, three times a week, for four weeks, an increase in BBS balance score was reported (43).

According to a meta-analysis of the effect of SET on balance in patients with stroke, SET treatment combined with conventional rehabilitation was found to be superior to conventional rehabilitation treatments, with increased degrees of BBS, BI, and FMA (Fugl Meyer assessment) in improving post-stroke balance function (43). Patients with neurological disorders, such as stroke, have mobility impairments, including balance and gait disturbances, which increase the risk of falls and affect their quality of life (44). Patients diagnosed with hemiplegia have asymmetric movements and reduced weight-bearing ability due to paralyzed limbs, which in turn affects balance and gait (45). Therefore, the ultimate goal is to improve the weight-bearing ability of paralyzed lower limbs in patients with stroke. Among various treatment methods, training using biofeedback is important to create normal movement patterns (46). In this study, core stabilization training using real-time feedback and a sling improved dynamic balance ability, which was consistent with a previous study that reported that imagined movements through visual and auditory stimuli affected weight shift and symmetry of patients with stroke by activating the motor area (47). Motion observation, which promotes the reconstruction of brain regions for motion, commands contributes to the formation of observed motor memory and promotes motor memory formation when combined with physical movement (48). It has been suggested that motion observation for patients with stroke is a more useful intervention method to improve motor function than simple task-oriented training (49). In this study, it was contemplated that training to control the trunk while providing visual and auditory feedback through the laser beam and verbal instructions of the therapist to help the patients concentrate had a positive effect on the patient’s motor memory.

In addition, core exercises using a sling have been reported to improve trunk stability and increase core muscle strength in a balanced manner, rotate the spine and vertebrae at the apex of left and right curvatures, correct the posture, and maintain the posture to further activate the core muscles (50).

### 4.2 The effects of gait ability

The gait pattern of patients with stroke not only shows speed reduction but also confusion regarding weight acceptance and transfer along with inefficient and unstable gait (51). Moreover, movement reportedly decreases in both temporal and spatial gait abilities (52). In this study, changes in gait before and after training were investigated using the TUG test and a spatiotemporal variable evaluation (GaitRite) of walking ability. In the TUG test, the RFCST group showed a significant reduction by 2.69 s from 22.70 s before the experiment to 20.01 s after the experiment, and the difference between the groups was found to be statistically significant (*p*<0.05). Among the gait variables, the walking speed of the RFCST group increased significantly by 5.57 cm/s from 37.05 cm/s before the experiment to 42.62 cm/s after the experiment. Moreover, the number of steps increased from 60.76 steps/min before the experiment to 64.68 steps/min after the experiment, demonstrating a significant increase of 3.92 steps /min. this difference between the groups was statistically significant (*p*<0.05).

In a study wherein balance and gait training was conducted using visual feedback for 24 patients with stroke for 30 min, three times a week for eight weeks, the experimental group showed improvements in TUG, BBS evaluation, gait speed, and walking distance compared to the control group (53). Moreover, in another study in which weight transfer and gait training was conducted using auditory feedback for 50 minutes, three times a week for six weeks in 24 patients with stroke, the experimental group showed improvement compared to the control group in terms of TUG and 10-meter walk test(10 MWT) evaluations (54). In this study, among the gait variables, the affected limb length significantly increased by 3.14 cm from 30.52 cm before the experiment to 33.66 cm after the experiment in the RFCST group, and the difference between the groups was found to be statistically significant (*p*<0.05).

In a previous study that showed similar results to ours, recumbent stepping, which requires trunk control, was performed with visual feedback by 11 patients with stroke over six months for 45 min at a time, three times a week for four weeks. The results confirmed that gait-related parameters improved in 5 Times Sit to Stand(5TSTS), Balance Master direction control and speed, and FUGL-Meyer lower extremity function. In the spatiotemporal gait parameter analysis using GaitRite, a significant increase in affected limb guarantee, affected lateral stride, and gait speed has been reported in the experimental group (*p*<0.05) (55). In addition, in a study in which core stabilization exercise was performed with real-time feedback training for 30 min a day for six weeks, the experimental group that performed core stabilization exercise through real-time feedback was significantly faster in the TUG test than the control group (*p*=0.042). Moreover, changes in stride length (*p*=0.021) and single support time (*p*=0.033) showed significantly greater improvements in the experimental group than in the control group (56).

Visual feedback applied in real-time reportedly affects gait by indicating the degree of motion of the participant at the same time as the motion, which aids in finding motion errors and aptly adjusting them (46). Moreover, it is also contemplated that mirror-like visual information is effective for motor learning as it conveys the participant’s previous and current movement information, thereby enabling them to realize errors and correct them (57). Motor learning is noticed to proceed through repeated corrections of motor commands based on motor errors (58). In this study, although the concentration and memory of patients with stroke deteriorated, the movement that the participant had to perform was provided with a laser beam to induce interest, and a sling was used to assist weight and focus on the trunk shape, while gravity was removed. This appeared to have increased the concentration.

To improve the ability to walk independently, it is important to promote the cognitive function that governs the walking process. The central nervous system is trained based on the motor learning theory of plasticity of movement, concentration of attention, and repetition of desired movements. Therefore, proprioceptive feedback is necessary for the efficient functioning of the central nervous system to restore and improve walking ability (55). Gait training provides real-time feedback on vertical toe displacement, a gait parameter that allows patients to adjust their toe spacing while walking on a treadmill, which reduces the probability of falling while the patient controls or changes the trajectory of their feet on their own. Gait ability can also be improved. Visual stimulation allows more focus on movement orientation, and through the visual and auditory information provided by the therapist, the patient can easily adjust and correct the trunk by themselves, thereby improving walking ability (59, 60).

A limitation of this study is that the small sample size and results obtained in a limited age range make it difficult to generalize the results for all patients with stroke. Moreover, it was also difficult to subdivide and apply the treatment plan or sequence tailored to each patient’s functional level. Depending on the quality of the disability, more or less fixed training is possible, and standardized treatment is impossible. Therefore, continuous research on individualized approaches that consider the patient’s disability criteria or conditions, various patterns, along family and social environmental influences in realistic treatment situations is needed. Furthermore, it is necessary to evaluate patients with neurological diseases from various aspects, considering that disorders appear in various areas (language and cognitive function, and emotional state), such as complex syndromes, and are not limited to motor dysfunction (61).

## 5. Conclusion

Balance issues commonly occur in patients after a stroke. Based on our findings, real-time feedback-based core stabilization training using a sling can be proposed as an effective treatment method for patients with stroke who have insufficient trunk stability. This treatment method can be actively used in clinical practice and would show good results.

## Institutional Review Board Statement

This study was conducted by the Declaration of Helsinki and was approved by the Institutional Review Board of Sahmyook University in Korea (SYUIRB2-1040781-A-N-012021059HR). The protocol of this trial was retrospectively registered in the Clinical Research Information Service of Korea (https://cris.nih.go.kr/cris/index/index.do; Registration Number: KCT0006552).

## Informed Consent Statement

Informed consent was obtained from all participants involved in the study.

## Data Availability Statement

Not applicable.

